# A Nation in Mourning: Prolonged Grief and Psychological Distress in the Israeli Public Amidst the Ongoing Hostage Crisis Following the October 7th Attack

**DOI:** 10.1101/2025.06.04.25328965

**Authors:** Yoav Groweiss, Carmel Blank, Yuval Neria, Yossi Levi-Belz

## Abstract

**Background:** The October 7, 2023, terrorist attack in Israel led to over 1,200 civilian deaths and the abduction of 251 individuals to Gaza. While prior studies have documented the psychological toll on directly affected populations, the broader emotional impact of the ongoing hostage crisis on the general public remains unclear. This study explored how concern for the hostages relates to psychological distress and functional impairment, conceptualizing this reaction as a form of collective prolonged grief.

**Methods:** A nationally representative sample of 515 Israeli adults completed self-report questionnaires at two timepoints: August 2023 (pre-attack) and May 2025. Measures assessed anxiety, depression, PTSD symptoms, cumulative stress, emotional burnout, and daily functioning. Concern for the hostages was rated on a 4-point scale. Prolonged Grief Disorder (PGD) symptoms were measured using an adapted PG-13 scale to reflect ambiguous loss. Multivariate analyses controlled for baseline distress and trauma exposure.

**Results:** Higher concern levels were significantly associated with elevated distress across all symptom domains. Nearly half (48.7%) of participants met criteria for probable PGD in relation to the hostage situation. These individuals exhibited significantly higher psychological symptoms and lower functional well-being—including poorer sleep, reduced concentration, and diminished optimism and hope.

**Conclusions:** Findings indicate that the prolonged hostage crisis constitutes a collective psychological burden marked by ambiguous loss and unresolved national trauma. This form of distress affects even those without direct exposure. Mental health efforts must extend beyond trauma-specific care to include interventions for symbolic and collective grief. National leadership plays a critical role in recognizing this suffering and restoring societal trust.

## Introduction

On October 7, 2023, Israel experienced an unprecedented and devastating terrorist attack orchestrated by the Hamas organization (Levi-Belz et al., 2024a). Approximately 1,200 civilians were killed, and 251 individuals—men, women, and children—were abducted from Israeli territory into the Gaza Strip, where they were held hostage by Hamas and the Palestinian Islamic Jihad (Vinograd & Kershner, 2023). The hostages represent a cross-section of Israeli society: infants, the elderly, civilians, soldiers, and foreign workers. Some were taken alive, while others were abducted posthumously (Levkovich et al., 2025). To date, eight hostages have been rescued through military operations, while sporadic agreements with Hamas have facilitated the cumulative release of 140 living hostages and the return of 49 bodies—some of whom are believed to have died in captivity. As of May 2025, 58 hostages remain in captivity, with at least 20 presumed alive (BBC News, 2025). The fate of these hostages has since become a source of profound national concern and societal discord, and testimonies from released hostages have further intensified these concerns (e.g., Levkovich et al., 2025).

It is well established that the October 7th terrorist attack and the subsequent war have had profound psychological effects (e.g., levels of anxiety, depression and posttraumatic symptoms) on the general population in Israel (Levi-Belz et al., 2024a; Feingold et al., 2024), as well as on civilians who experienced direct exposure to the attack (Levi-Belz et al., 2025a), civilians who experienced loss of loved one (Levi-Belz et al., 2025b) and evacuation (Amsalem et al., 2025) following the attack and the subsequent war (Levi-Belz et al., 2025c). However, the emotional impact of the ongoing public concern surrounding the hostage crisis remains largely unexplored. In particular, it is unclear whether—and to what extent—this collective concerns regarding the hostages’ situation contributes to elevated psychological symptoms. Clarifying this issue is essential for understanding how unresolved national trauma and symbolic, indirect threats shape public mental health and societal resilience.

Despite the centrality of the hostage crisis in Israeli public discourse, empirical data on civilians’ emotional responses to the ongoing captivity remain limited. For example, Yehene et al. (2024) found that 93.1% of Israelis reported being preoccupied with the hostages’ well-being, and many expressed only low to moderate confidence in their eventual return—highlighting the psychological burden of sustained uncertainty. Public concern is also reflected in widespread civic engagement: ongoing rallies, vigils, media campaigns, and visible symbols such as yellow ribbons and posters have become part of the national landscape. In this context, perceived governmental betrayal—stemming from the state’s failure to prevent the abductions or to secure the hostages’ release—has also been identified as a factor that may intensify psychological distress (Levi-Belz et al., 2024b). Taken together, these trends underscore the importance of understanding how prolonged national crises—and the public’s sustained emotional engagement with them—may affect the mental health of an entire society.

### Worry as a Psychological Construct

Worry is defined as a chain of future-oriented thoughts and images concerning uncertain and potentially negative outcomes (Sibrava & Borkovec, 2006). While worry is a normative human experience, it becomes pathological when it is excessive, uncontrollable, and distressing, leading to functional impairment (Barlow, 2002). Pathological worry is a hallmark feature of generalized anxiety disorder (GAD; APA, 2022) but is also implicated in other psychopathologies, including depression (Yook et al., 2010; Bentley et al., 2016) and PTSD (Tull et al., 2011). Studies have demonstrated that worry is positively associated with anxiety (Casey et al., 2004), depression (Starcevic, 1995; Buck et al., 2008), and somatic stress symptoms (Brosschot & van der Doef, 2006). Furthermore, worry has been linked to emotional avoidance, depletion of coping resources, and burnout (Shin et al., 2014), including in contexts of prolonged national crises such as war (Elhadi et al., 2020; Tsybuliak et al., 2023). Given the high levels of public concern reported in Israel (Yehene et al., 2024), it is plausible to hypothesize that worry over the hostages’ fate contributes to heightened psychological distress such as anxiety, depression, posttraumatic stress symptoms and burnout.

### Prolonged Grief in the Context of Collective Trauma

A further dimension of the psychological impact relates to prolonged grief disorder (PGD). PGD is defined as persistent and pervasive grief that lasts beyond culturally accepted norms (typically over 12 months in DSM-5-TR and six months in ICD-11), accompanied by intense yearning, preoccupation with the loss, and significant emotional pain, leading to functional impairment (APA, 2022; WHO, 2000). PGD has been linked to elevated levels of anxiety, depression, PTSD, and impairments in daily functioning (Prigerson et al., 2021; APA, 2022). While grief typically follows death, the concept of ambiguous loss—loss without closure or certainty—may also elicit similar psychological responses (Boss, 1999; Lundorff et al., 2017). Research has demonstrated that ambiguous loss, such as disappearance without confirmation of death, is associated with more severe PGD symptoms compared to confirmed losses (Lundorff et al., 2017).

In the context of the hostage crisis, the ambiguity surrounding the hostages’ fates, coupled with their symbolic representation in Israeli society, may function as a form of collective ambiguous loss (Yehene et al., 2024). The inability to mourn, the persistent exposure to the hostages’ stories in the media, and the collective struggle to balance hope and despair have likely created a unique form of grief that pervades Israeli society. The ongoing uncertainty about the hostages’ status—alive or dead—profoundly impacts individuals’ capacity to move forward, echoing the dynamics of PGD. Thus, it is plausible to suggest that individuals experiencing higher levels of prolonged grief symptoms related to the hostages will report greater psychological distress (including anxiety, depression, PTSD, stress, and burnout) and impaired functioning in daily life.

### The Present Study

The ongoing captivity of Israeli hostages following the October 7th terrorist attack constitutes a unique form of collective, ambiguous loss that continues to shape public discourse and emotional life in Israel. As reviewed above, while research has documented elevated distress among individuals directly exposed to the attack, bereaved families, and evacuees (e.g. Levi-Belz et al., 2025a; Amsalem et al., 2025), far less is known about the broader psychological toll of the hostage crisis on the general public—particularly when the loss is symbolic, unresolved, and deeply national in nature. The current study aimed to fill this gap by examining the psychological and functional correlates of public concern regarding the hostages. Specifically, we focused on two key psychological constructs that may underlie this distress: **worry**, a future-oriented cognitive process, and **prolonged grief**, particularly as it may manifest in response to ambiguous or unresolved loss at the collective level.

To address these aims, we leveraged data from a prospective longitudinal study conducted on a nationally representative sample of Israeli citizens (e.g., Levi-Belz et al., 2024a). The current analysis focuses on the most recent wave of data collection, conducted in April–May 2025, approximately one and a half years after the October 7th terrorist attack. Importantly, all participants had previously completed baseline assessments of psychological distress in late August 2023, roughly five weeks prior to the attack. This pre-crisis dataset allowed us to statistically adjust for individual differences in pre-existing mental health, thereby enabling a more precise examination of the psychological impact associated with the ongoing hostage crisis. Our hypotheses were:

**H1:** Greater concern about the fate of the hostages will be significantly associated with elevated psychological distress, including higher levels of anxiety, depression, posttraumatic stress symptoms, cumulative stress, and burnout—even after accounting for baseline mental health levels.

**H2:** Participants meeting criteria for probable Prolonged Grief Disorder (PGD) following the hostages’ situation will exhibit significantly higher psychological distress across all measured domains (anxiety, depression, PTSD symptoms, stress, and burnout), compared to participants without PGD.

**H3:** Individuals with probable PGD will demonstrate significantly lower functional well-being, including poorer daily functioning, reduced sleep quality and concentration, and diminished levels of optimism, inner calm, and hope for the future compared to participants without PGD.

## Methods

### Participants and Design

The final study sample consisted of 515 Israeli adults who completed both waves of data collection. Eligibility criteria required participants to be Israeli citizens residing in Israel at the time of the October 7th terrorist attack.

Data were collected at two time points. The first wave (T1) was conducted in August 2023, approximately one and a half months prior to the attack, and served as a baseline assessment of psychological distress. The second wave (T2) was conducted in May 2025, approximately 18 months after the attack, during the ongoing war. A total of 908 participants completed the baseline survey at T1, and 515 of them responded again at T2, yielding a follow-up response rate of 56.7%.

Comparative attrition analyses revealed no significant differences in key sociodemographic characteristics (age, gender, SES) or baseline symptom levels between those who completed both waves and those lost to follow-up, supporting the validity of the longitudinal sample.

### Measures

#### International Trauma Questionnaire (ITQ; Cloitre et al., 2018)

Posttraumatic stress symptoms were assessed using the nine PTSD-related items of the ITQ, based on ICD-11 criteria. The scale includes six items representing re-experiencing, avoidance, and threat, rated on a 5-point Likert scale (0–4), and three additional items assessing functional impairment. The ITQ has demonstrated strong psychometric properties (Shevlin et al., 2018) and showed excellent internal consistency in this study (T1 α = .86; T2 α= .87).

#### Patient Health Questionnaire-2 (PHQ-2; Kroenke et al., 2003)

The PHQ-2 is a brief, validated depression screener assessing the two core DSM-5 symptoms of major depression: anhedonia and depressed mood. Items are rated on a 4-point Likert scale (0–3). This tool has been shown to have high sensitivity and specificity and yielded solid internal consistency in the present sample (T1 α = .82; T2 α= .86).

#### Generalized Anxiety Disorder-2 (GAD-2; Kroenke et al., 2007)

Anxiety symptoms were measured using the GAD-2, a validated 2-item scale based on the GAD-7. Participants rated the frequency of symptoms such as nervousness and uncontrollable worry over the past two weeks. Internal consistency was high across both waves (T1 α = .86; T2 α= .87).

#### Perceived Stress Scale (PSS; Cohen, Kamarck, & Mermelstein, 1983)

Perceived stress was assessed using the PSS, a widely used 10-item self-report scale that measures the extent to which individuals appraise situations in their lives as stressful, unpredictable, and overwhelming. Items (e.g., “In the last month, how often have you felt difficulties were piling up so high that you could not overcome them?”) are rated on a 5-point Likert scale ranging from 0 (“never”) to 4 (“very often”). Mean scores are calculated by summing item responses, with higher scores indicating greater perceived stress. The PSS has demonstrated strong validity and reliability across populations and yielded high internal consistency in the current study (T2 α =.91).

#### Maslach Burnout Inventory – Emotional Exhaustion (MBI; Maslach & Jackson, 1981)

Emotional burnout was measured using the emotional exhaustion subscale of the MBI, consisting of 9 items assessing fatigue, emotional depletion, and reduced energy (e.g., “I feel emotionally exhausted because of my work”). Items were rated on a 5-point Likert scale from 0 (“never”) to 4 (“daily”), and a mean score was computed. This subscale reflects both physical and emotional aspects of burnout and demonstrated excellent internal consistency in this study (α = .91–.92).

#### Functional Impairment

Subjective functional status was assessed using a brief, five-item self-report scale developed for the current study, designed to capture perceived changes in day-to-day functioning relative to the pre-war period. The scale was conceptually informed by established functional and well-being measures such as the Functional Status Questionnaire (FSQ; Jette et al., 1986) and the WHOQOL-BREF (WHO, 1998) and was adapted to capture the unique impact of prolonged national trauma. Each item was rated on a 7-point Likert scale ranging from 1 (“Much worse than before the war”) to 7 (“Much better than before the war”), with 4 indicating no change. Higher scores reflected improved functioning compared to pre-war status, while lower scores indicated functional decline. The five items assessed core domains of civilian life including general daily functioning, sleep quality, concentration, emotional calm, and future outlook.

#### Prolonged Grief – Prolonged Grief Disorder Scale (PG-13; Prigerson et al., 2009)

Prolonged grief was assessed using the PG-13, a validated 13-item self-report scale developed to identify individuals meeting criteria for Prolonged Grief Disorder (PGD). In the present study, the items were adapted to reflect the context of ambiguous loss associated with the hostages in Gaza, rather than the death of a specific individual. For example, the item “In the past month, how often have you felt intense emotional pain, sorrow, or pangs of grief about the death?” was rephrased as: “In the past month, how often have you felt intense emotional pain, sorrow, or grief regarding the hostages in Gaza?”

Participants responded on a 5-point Likert scale (1 = not at all to 5 = several times a day/overwhelmingly), and a total symptom severity score was calculated. A validated cutoff score of ≥30 (**Prigerson et al., 2009**) was used to classify individuals as meeting the threshold for probable PGD. This dichotomous variable (probable PGD vs. non-PGD) was used as a key grouping variable in all analyses. Internal consistency for the adapted scale was high (α = .86).

### Sociodemographic and Trauma-Related Characteristics

Participants provided background information on key sociodemographic variables, including age, gender, and socioeconomic status (SES). In addition, trauma-related exposure was assessed across three primary domains. First, direct exposure to the October 7th terrorist attack was defined as being physically present in towns or communities within the Gaza envelope during the assault. Second, bereavement was defined as the loss of a close family member or loved one who was murdered in the attack. Third, participants reported whether they had been called up for active reserve military duty during the ongoing war, and whether they served in a combat role for more than 30 consecutive days.

As part of a broader set of items assessing personal, economic, and national concerns related to the war, participants were asked to rate their level of worry about the Israeli government’s handling of the hostage crisis. Responses were recorded on a 4-point Likert scale ranging from 1 (“Not at all worried”) to 4 (“Very worried”). This item was analyzed as an independent variable reflecting public concern regarding an unresolved and emotionally salient national issue.

### Procedures

Study participants were recruited online via Panel4All, a professional survey company that maintains a probability-based panel of approximately 100,000 Israeli respondents. The sampling procedure was quota-based, ensuring representation of the general Israeli population according to national census data on age, gender, ethnicity, and socioeconomic status. Participants received monetary compensation from the survey company for their time.

The current analyses are based on data collected at two time points. The first assessment (T1) took place approximately one month prior to the October 7th terrorist attack and served as a baseline measure of psychological distress. The second assessment (T2) was conducted about 18 months later, during the ongoing war, and included measures of psychological symptoms, functional impairment, and attitudes toward the hostage crisis. At both time points, participants received a link to the online questionnaire via Qualtrics. Participation was voluntary, and all respondents provided informed consent before completing the survey. Anonymity and confidentiality were assured, and participants were informed of their right to withdraw at any point. The study was approved by the ethics committee of the Ruppin Academic Center (Protocol No. 175/2023).

### Data Analysis

A series of multivariate analyses of covariance (MANCOVA) were conducted to examine the psychological and functional correlates of (1) levels of concern about the hostages and (2) probable Prolonged Grief Disorder (PGD) related to the hostage situation. Each predictor was entered as a fixed independent factor in separate models. Dependent variables included five psychological distress indicators (anxiety, depression, PTSD symptoms, cumulative stress, and burnout) and six functional outcomes (daily functioning, sleep quality, concentration, optimism, inner calm, and hope for the future).

Participants’ baseline psychological distress levels – Anxiety, depression and PTSD symptoms, assessed at T1 (August 2023, prior to the October 7th attack), were included as covariates in all psychological models to control for pre-existing symptom severity. Additional covariates included age, gender, socioeconomic status, and trauma-related exposure variables (direct exposure to the attack and bereavement).

Assumptions of MANCOVA—including multivariate normality, linearity, homogeneity of variance-covariance matrices (Box’s M test, p > .001), and absence of multicollinearity—were tested and met. Where significant multivariate effects emerged, follow-up univariate ANOVAs were conducted to examine specific group differences. Pairwise comparisons were performed using the least significant difference (LSD) method. Partial eta squared (η²) values were computed to estimate effect sizes. All analyses were conducted using SPSS version 28, with statistical significance set at *p* < .05.

## Results

### Participants’ Demographics and Hostage-Related Concern and Grief

The sample’s demographics and October 7th attack-related variables are presented in Table 1. The final study sample comprised 515 participants, aged between 18 and 85 years (M = 41.02, SD = 13.79), with an approximately equal gender distribution (n = 259 women; 50.3%).

Participants were asked to report their current level of concern regarding the hostage crisis, as part of a broader set of items addressing war-related worries (e.g., personal, economic, and national concerns). Responses were recorded on a 4-point Likert scale ranging from 1 (“Not at all worried”) to 4 (“Very worried”). Among the participants, 7.0% were “Not at all worried,” 17.5% were “Not worried,” 29.9% were “Worried,” and 45.6%—nearly half the sample—were “Very worried”.

With respect to prolonged grief related to the hostage situation, we examine the levels of probable Prolonged Grief Disorder (PGD). Of these, 48.7% (n = 246) met criteria for probable while 51.3% (n = 259) did not. This near-equal distribution highlights the deep emotional toll and ongoing psychological relevance of the hostage crisis within the general Israeli population.

### Differences in distress levels according to the levels of worries regarding the hostages

To examine the psychological implications of public concern regarding the government’s handling of the hostage situation, we conducted a MANCOVA analysis. The model tested the effect of worry about the hostages (with four levels: “Not at all worried,” “Not worried,” “Worried,” and “Very worried”) on five psychological outcomes: anxiety, depression, PTSD symptoms, stress, and emotional burnout. Importantly, participants’ baseline psychological distress levels, measured prior to the October 7th attack (T1), were entered into the model as a covariate to control for pre-existing mental health differences.

Multivariate analysis revealed a significant effect of worry level on the combined set of psychological outcomes [Pillai’s Trace = .090, F(15, 1497) = 3.094, p < .001, η² = .030]. Univariate tests showed significant effects across all outcomes: anxiety [F(3, 501) = 5.73, p < .001, η² = .033], depression [F(3, 501) = 6.79, p < .001, η² = .039], PTSD symptoms [F(3, 501) = 5.72, p < .001, η² = .033], stress [F(3, 501) = 10.76, p < .001, η² = .061], and emotional burnout [F(3, 501) = 10.03, p < .001, η² = .057]. As it can be seen on Table 2, post-hoc pairwise comparisons indicated that participants who reported being “very worried” about the hostages consistently exhibited higher levels of distress compared to those with lower worry levels. For instance, anxiety scores among the “very worried” group were M = 1.86 (SD = 1.79), compared to M = 0.97 (SD = 1.78) among those “not at all worried.”

**Table 2.**
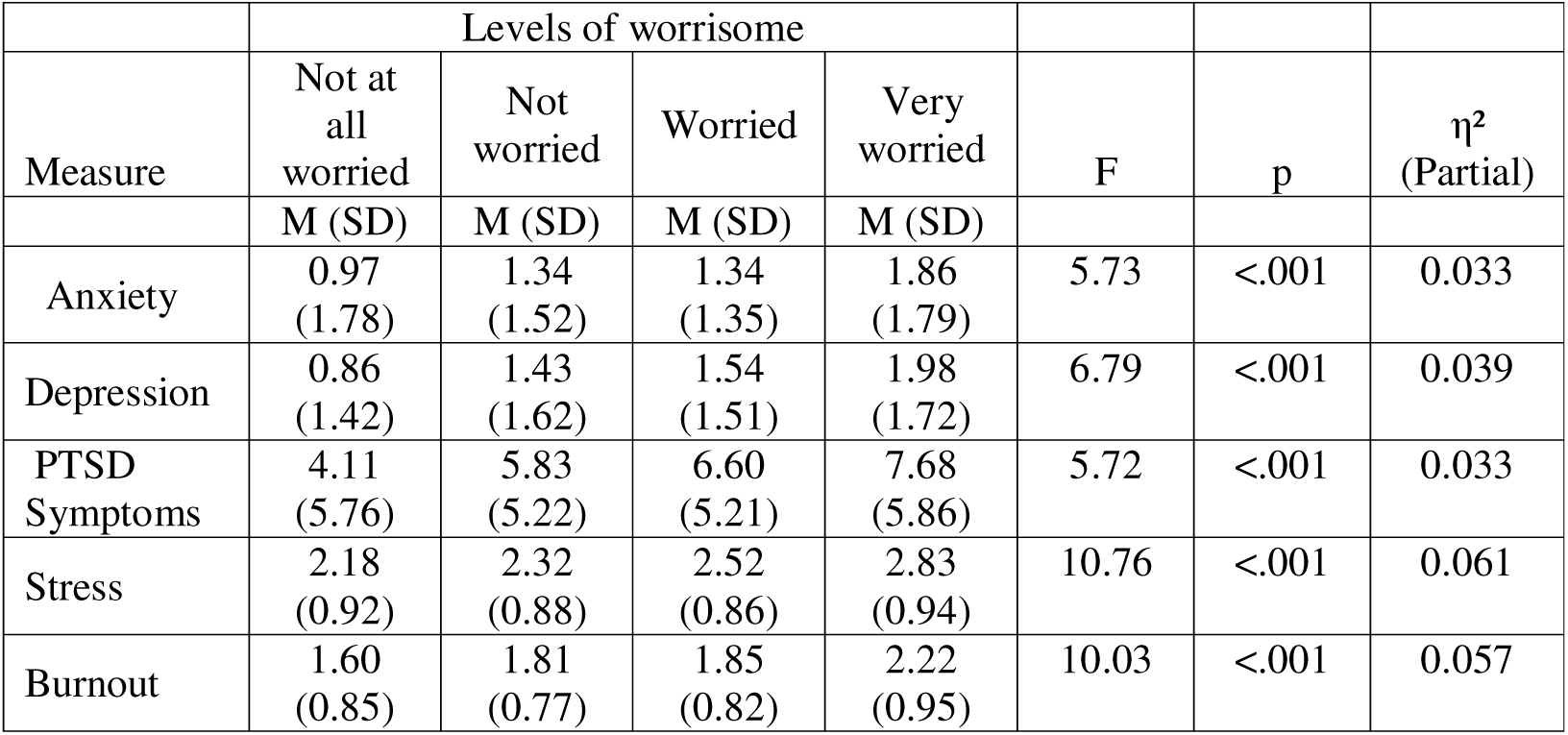
Differences in distress levels according to the levels of worries regarding the hostages.

### Differences in distress levels according to Probable Prolonged Grief Disorder (PGD) regarding the hostages

To evaluate the psychological consequences of prolonged grief in the context of the hostage crisis, a MANCOVA analysis was conducted comparing participants who met the criteria for probable Prolonged Grief Disorder (PGD; cutoff ≥ 30) with those who did not. The analysis included five psychological outcomes: anxiety, depression, PTSD symptoms, cumulative stress, and burnout. Participants’ baseline psychological distress levels(T1) were used as a covariate to control for pre-existing mental health differences.

Multivariate analysis revealed a significant overall effect of PGD status on the combined outcomes [Pillai’s Trace = .153, F(5, 499) = 18.03, p < .001, η² = .153]. As it can be seen on Table 3, univariate tests revealed significant group differences for each outcome, as participants meeting the probable PGD criteria consistently exhibited substantially higher symptom levels in all measures: anxiety [F(1, 503) = 60.01, p < .001, η² = .107], depression [F(1, 503) = 69.15, p < .001, η² = .121], PTSD symptoms [F(1, 503) = 56.94, p < .001, η² = .102], stress [F(1, 503) = 43.67, p < .001, η² = .080], and emotional burnout [F(1, 503) = 71.68, p < .001, η² = .125].. For instance, their mean anxiety level was M = 2.10 (SD = 1.71), compared to M = 1.03 (SD = 1.40) among non-PGD participants.

**Table 3.** Differences distress levels according to Probable Prolonged Grief Disorder (PGD) regarding the hostages.

| Measure | No PGD | With probable PGD | F | p | $\eta^2$ (Partial) |
| --- | --- | --- | --- | --- | --- |
| Anxiety | 1.03 (1.40) | 2.10 (1.71) | 60.01 | <.001 | 0.107 |
| Depression | 1.12 (1.39) | 2.26 (1.70) | 69.15 | <.001 | 0.121 |
| PTSD Symptoms | 5.04 (5.04) | 8.63 (5.65) | 56.94 | <.001 | 0.102 |
| Stress | 2.34 (0.89) | 2.87 (0.90) | 43.67 | <.001 | 0.080 |
| Burnout | 1.69 (0.76) | 2.32 (0.92) | 71.68 | <.001 | 0.125 |

### Differences functional levels according to Probable Prolonged Grief Disorder (PGD) regarding the hostages

To further explore the functional impact of Prolonged Grief Disorder (PGD) in the context of the ongoing hostage crisis, a MANCOVA analysis was conducted comparing participants with probable PGD to those without. The model included six self-reported functional outcomes: daily functioning, sleep quality, concentration ability, optimism, inner calm, and future hope.

Multivariate analysis revealed a significant overall effect of PGD status on functional outcomes [Pillai’s Trace = .041, F(6, 465) = 3.303, p = .003, η² = .041]. As it can be seen on Table 4, univariate analyses demonstrated that participants meeting criteria for probable PGD consistently reported significantly poorer functioning across all domains: daily functioning [F(1, 470) = 10.74, p = .001, η² = .022], sleep quality [F(1, 470) = 12.95, p < .001, η² = .027], concentration [F(1, 470) = 12.35, p < .001, η² = .026], optimism [F(1, 470) = 14.27, p < .001, η² = .029], inner calm [F(1, 470) = 16.69, p < .001, η² = .034], and future hope [F(1, 470) = 14.63, p < .001, η² = .030].

**Table 4.**
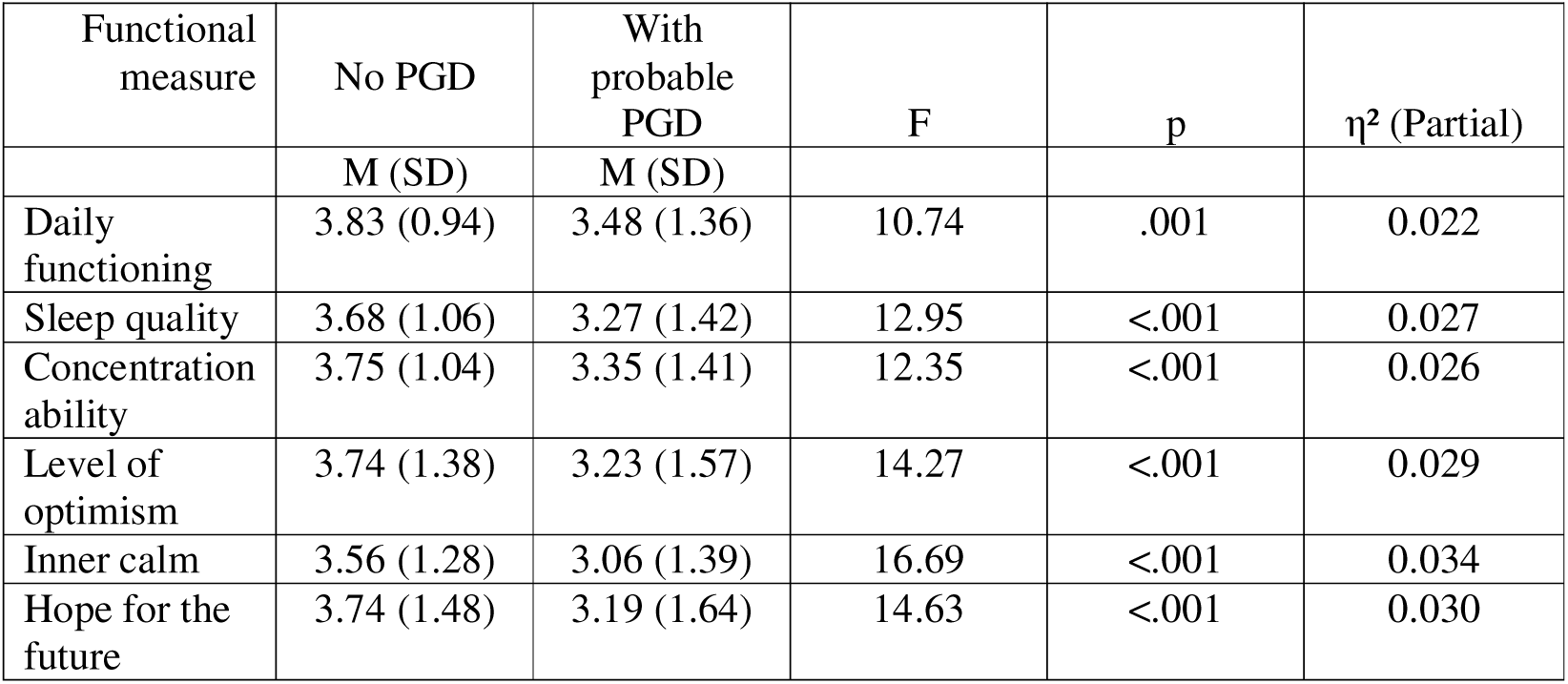
Differences functional levels according to Probable Prolonged Grief Disorder (PGD) regarding the hostages.

| Functional measure | No PGD | With probable PGD | F | p | $\eta^2$ (Partial) |
| --- | --- | --- | --- | --- | --- |
|  | M (SD) | M (SD) |  |  |  |
| Daily functioning | 3.83 (0.94) | 3.48 (1.36) | 10.74 | .001 | 0.022 |
| Sleep quality | 3.68 (1.06) | 3.27 (1.42) | 12.95 | <.001 | 0.027 |
| Concentration ability | 3.75 (1.04) | 3.35 (1.41) | 12.35 | <.001 | 0.026 |
| Level of optimism | 3.74 (1.38) | 3.23 (1.57) | 14.27 | <.001 | 0.029 |
| Inner calm | 3.56 (1.28) | 3.06 (1.39) | 16.69 | <.001 | 0.034 |
| Hope for the future | 3.74 (1.48) | 3.19 (1.64) | 14.63 | <.001 | 0.030 |

## Discussion

This study aimed to explore the psychological and functional consequences of the ongoing hostage crisis in Israel, following the prolonged and emotionally charged crisis of the October 7^th^ attack and the subsequent war. Specifically, we leveraged a nationwide prospective study designed to capture mental health dynamics in the aftermath of the October 7th attack (see details at Levi-Belz at al., 2025). Within this framework, we investigated how varying levels of worry regarding the fate of hostages held in Gaza relate to symptoms of anxiety, depression, posttraumatic stress, stress, emotional burnout, as well as of daily functioning impairment beyond baseline levels of distress measured one an a half months before the attack (T1). By integrating both symptom-related and functional outcomes, this investigation may offer a comprehensive view of the public mental health burden posed by a hostages crises in Israel.

Our findings highlight the profound psychological and functional disruption experienced by the Israeli citizens who are worried regarding the hostages situation. The results indicated participants with higher levels of concern for the hostages’ fate reported significantly higher levels of psychological distress, including elevated anxiety, depression, PTSD symptoms, stress, and burnout. These results are consistent with prior research demonstrating the strong link between worry and mental health difficulties (Casey et al., 2004; Yook et al., 2010; Bentley et al., 2016; Tull et al., 2011). Moreover, our results indicated that participants meeting probable PGD criteria in regard to the ambiguous loss following the hostages’ situation exhibited significantly higher levels of anxiety, depression, PTSD symptoms, stress, and emotional burnout as well as higher levels of daily functioning impairment compared to those without PGD.

These results align with existing literature on the detrimental impact of PGD on mental health (e.g., Prigerson et al., 2021), reinforcing the conceptualization of the public’s ongoing concern for the hostages as a form of pathological grief. Additionally, the findings regarding the lower daily functioning impairment among participants with probable PGD regarding the hostages’ situation may emphasize the far-reaching impact of complicated grief, extending beyond emotional suffering to substantial disruptions in basic life functioning—effects that persist months after the initial traumatic events (Kristensen et al., 2014; Nielsen et al., 2020).

It is important to note that participants in this study did not personally know the hostages who remain in captivity in Gaza. This concern is anchored in national identification and is continually intensified by extensive continuous media coverage, public discourse, and the unresolved nature of the crisis (Yehene et al., 2024). Based on the high proportion of citizens reporting significant concern and symptoms consistent with PGD in response to the hostages crisis, it is plausible to suggest that the distress observed in this study does not stem from direct relational loss. Rather, it reflects a broader collective experience—one rooted in national identification, perceived moral obligation, and a shared sense of helplessness in the face of unresolved trauma (Hirschberger, 2018). This phenomenon is likely amplified by continuous media exposure and ongoing governmental inaction, which together reinforce the perception of collective abandonment (Yehene et al., 2024) and moral injury of betrayal (Levi-Belz et al., 2024b; Litz and Walker, 2025).

In general, our findings provide empirical support for the view that the hostage crisis constitutes a form of *collective ambiguous loss* (Boss, 2004) embedded within a traumatic national narrative (Hirschberger, 2018). This pattern of loss is characterized by the inability to mourn, the absence of closure, the continuous presence of reminders of the hostages’ absence, and the persistent uncertainty regarding their fate (Boss, 2004; Yehene et al., 2024) - highlight the psychological costs of prolonged exposure to unresolved, morally charged events.

Importantly, our study extends these previous findings by highlights the collective context and emphasizing how ongoing, unresolved national crises may generate widespread psychological distress that transcends personal vulnerability. Taken together, these findings underscore the urgent need to recognize ambiguous loss not merely as an individual phenomenon but as a collective psychological burden such as in the hostages’ situation in Israel —one that shapes public mental health, challenges societal resilience, and demands context-sensitive therapeutic and policy responses during prolonged national crises.

Several limitations warrant consideration while interpreting the study results. First, the reliance on self-report measures introduces the possibility of response biases, including recall inaccuracies and social desirability effects. A related important limitation is the novel use of the PG-13 scale (Prigerson et al., 2009) which was adapted to capture the specific features of the hostage crisis—namely, the absence of certainty and closure – and not bereavement regarding a loved one death. Future studies should further differentiate the conceptual and clinical boundaries between ambiguous loss and prolonged grief, particularly in contexts where no definitive or finite loss has occurred (see also Manevich et al., 2023). Second, while we took into account the participants baseline levels of distress (at T1), unmeasured variables—such as ongoing exposure to related traumatic events engagement in psychological treatment, or social support characteristics may have influenced the observed outcomes. Future studies should investigate potential moderators, including demographic and cultural factors, previous trauma exposure, and individual coping strategies, to refine the understanding of how collective ambiguous loss manifests across populations. Finally, the study’s focus on the Israeli context, with its unique sociopolitical and cultural dimensions, may limit the generalizability of findings to other populations experiencing collective trauma.

## Conclusions and Implications

This study offers critical insights into the psychological toll of the ongoing hostage crisis on the Israeli public, highlighting it as a unique and unprecedented form of collective trauma.

Our findings demonstrate that persistent public concern for the hostages is associated with heightened levels of anxiety, depression, PTSD symptoms, stress, and emotional burnout. Furthermore, the uncertainty surrounding the hostages’ fate evokes a grief response that aligns with the clinical features of PGD, including profound emotional distress and substantial daily functional impairment. By framing the public’s reaction as a collective grief response—rooted not only in the death of loved one citizens but also in a broader lack of trust in a leaders perceived as failing to protect its people—this study underscores the complex layers of psychological distress in the context of ongoing national trauma.

These findings highlight a critical need to broaden our understanding of psychological distress in the context of national trauma. Alongside direct exposure (Levi-Belz et al., 2025b) and the personal loss of loved ones during the October 7th attack and ensuing war (Levi-Belz et al., 2025c), many citizens are profoundly affected by the ongoing hostage crisis. This prolonged uncertainty has led to widespread emotional pain and functional difficulties, even among those not directly exposed. Mental health professionals must therefore assess symptoms not only among those who experienced direct trauma, but also among individuals suffering from the enduring, unresolved nature of the hostage situation. Targeted interventions addressing ambiguous loss are essential to support those grappling with this collective burden.

Beyond the clinical implications, our findings highlight a critical role for national leadership in times of prolonged collective trauma. The psychological toll of the hostage crisis is not confined to those with direct exposure or loss; it reverberates through society, affecting countless individuals who experience ongoing distress, uncertainty, and functional impairment. In this context, leadership must not only act, but also be seen to act—with visibility, empathy, and clarity. Recognizing the public’s emotional burden and validating their grief is not a symbolic gesture—it is a vital component of national resilience. When leaders acknowledge suffering, communicate transparently, and demonstrate genuine commitment to resolving the crisis, they restore a sense of containment and reforge the frayed social contract. Our study suggests that such visibility and moral responsiveness are not merely political necessities but mental health imperatives. The failure to address this dimension risks deepening public disillusionment and eroding psychological cohesion. As such, healing from this ongoing trauma requires not only therapeutic intervention but also trustworthy, responsive leadership that signals: *we see your pain, and we are doing everything in our power to end it*.

## Data Availability

All data produced in the present study are available upon reasonable request to the authors

